# Using machine learning to discover correlations in MIMIC high-cadence data sets

**DOI:** 10.1101/2025.09.01.25334873

**Authors:** Viktor T. Toth, Samy A. Mahmoud

## Abstract

Using the MIMIC-IV clinical database and, specifically, the related preliminary MIMIC-IV waveform data release, we endeavored to create a machine learning tool in the form of a gated recurrent unit (GRU) neural network. Our goal was to study the possibility that intrusive measurements of vital signs, such as blood pressure, might be reliably predicted from parallel time series data sets of non-intrusive measurements of other vital signs, such as heart and respiratory rates. Relying on non-intrusive measurements to alert health care professionals of potentially life-threatening conditions in a timely manner can be life-saving in certain situations. To this end we developed and implemented a custom GRU solution in the form of software that can be run in a Web browser to analyze the high cadence numerical data sets of the MIMIC-IV waveform release. Despite the limitations of this data set, such as the small number of records, inconsistencies in what has been recorded, and often, noisy or incomplete data, we were able to obtain potentially promising results.

## I. Introduction

Version 2.4 of the Medical Information Mart for Intensive Care IV (MIMIC-IV) [1], [2], [3] database is the latest version of emergency room (ER) records collected over a number of years from several million emergency room events. The records are anonymized, allowing for the use of the data for analysis and research.

The MIMIC-IV database has several major components. These include anonymized patient information, low cadence time series data (e.g., intermittent blood pressure measurements) as well as high cadence data sets [4], originally recorded as waveform data, from which high-cadence time series data sets were extracted. The data records cover patients’ ER visits in full or in part.

Our main interest that led us to exploring the content and utility of the MIMIC-IV data set was the possibility that there exist correlations that can be used as predictors of intrusive measurements based on non-intrusive (or less intrusive) observables. As an example, if it is possible to reliably predict, from observations such as heart rate or electrocardiograms, significant changes in a patient’s blood pressure, this will make it feasible to alert health care workers to a potentially life-threatening condition that requires intervention in a timely manner, without subjecting the patient to recurrent, disruptive and often irritating blood pressure measurements using intrusive methods.

The current version (0.1.0) of the MIMIC-IV waveform database is preliminary, with data from fewer than 200 patients. Nonetheless, even this limited data set is already representative of the kind of information that we can utilize in our analysis.

Our goal was to establish a suitable machine learning strategy to analyze this data and, if possible, build an initial tool set that can demonstrate the utility of a machine learning approach in this analysis. In the following, we introduce a machine learning model that we developed for this purpose and report on our progress, while noting the availability of the code that we have developed to date.

In Section II, we introduce the basic concepts behind machine learning analysis of time series data, specifically the use of a gated recurrent unit (GRU) network for this purpose. The MIMIC-IV data release and the associated waveform data release are described in Section III. In Section IV we discuss our preliminary results. Some conclusions and possible directions of further work are presented in Section V.

## II. Analyzing time series data using a gated recurrent unit neural net

Given a data set that consists of several parallel streams of time series data, our aim is to discover two types of relationships:

1. Autocorrelation within a time series, which can help predict the future behavior of the time series based on past data; and
2. Correlation between the time series, which can help predict the values of one time series data from values from one or several other time series data.

In principle, both goals described above may be realizable using appropriate machine learning tools. The two approaches may also appear in combination, meaning that correlation and autocorrelation within a set of several time series may help predict the future behavior of the same set of time series.

Prediction of future behavior based on past data, however, is beyond the scope of our present investigation. Our goal is limited to deploying a neural network with the ability to discover nonlinear correlations between parallel time series data sets, thereby allowing us to predict the behavior of one time series from a selection of other time series. As an example, our approach may allow us to reliably predict blood pressure data using parallel time series data from non-intrusive or less intrusive measurements, such as heart rate or electrocardiogram (ECG) parameters.

There are a variety of approaches to analyze time series data [5], [6], [7]. A basic approach is to use a Recurrent Neural Network [8], [9] (RNN – see Fig. 1). The input layer of an RNN consists of cells that receive one block of input data. The cells predict output by applying an activation function to the weighted sum of their input and the output of the preceding cell in the time series sequence. Multiple (hidden) layers with a variable number of cells may be present with their respective outputs and inputs connected.

**Fig. 1.**
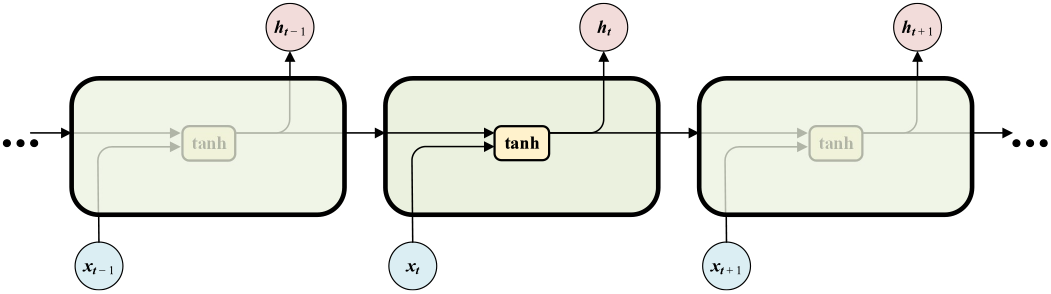
Schematic of a layer in a recurrent neural network (RNN). The network maps its input *x_t_*to its hidden-layer output *h*_*t*_ using internal weights, the output constrained by an activation function, typically the hyperbolic tangent.

Specifically, the output (hidden state) of an RNN cell is determined by an expression such as

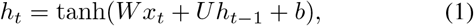

where *W* and *U* are learned weights and *b* is a learned bias. The activation function is typically the hyperbolic tangent. The model may also incorporate learned biases in addition to the learned weights.

Note that the inputs to an RNN, i.e., the quantities *x*_*t*_, may be vector-valued quantities. Similarly, the hidden-state output of an RNN cell may also be vector-valued. In this case, we may write

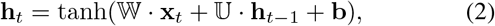

indicating also that in this case, 𝕎 and 𝕌 are matrix-valued. The dimensionality of **x**_*t*_ and **h**_*t*_ may differ.

An RNN may include multiple layers. The input dimensionality of each layer must be the same as the hidden state dimensionality of the preceding layer. The final output of the network is further weighted and biased, as in

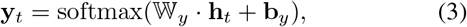

where the softmax function is a normalization function, e.g., normalizing the output values to sum up to 1.

The number of cells in a layer determine the length of the data sequence that the layer can process. The same network, with the same layers, weights and biases is reused for subsequent sequences. An RNN implementation is considered *stateful* if the input hidden state *h*_0_ used for its first cell is, in fact, the final hidden state output *h*_*n*_ produced by processing the preceding sequence. This approach allows an RNN to process time series data of arbitrary length. The sequence length that characterizes the dimension of the RNN is an important hyperparameter that must be adjusted to yield best results for the data set under analysis.

When the network is trained, weight matrices and bias vectors are computed by means of backpropagation: this involves defining a loss function that characterizes the accuracy with which the network can reproduce desired results, computing partial derivatives of the loss function with respect to the weights, and then updating the weights to minimize the loss function using a steepest descent or related method.

Unfortunately, simple RNNs have known shortcomings, including unstable gradients (resulting in numerical values that either diverge of fail to converge on a useful result) and the inability to effectively capture long-term relationships.

These shortcomings of RNNs are addressed by more sophisticated solutions. Two particular popular approaches are Long Short-Term Memory (LSTM) and Gated Recurrent Unit (GRU) solutions [10], [11]. By incorporating additional feed-back mechanisms, these solutions result in neural networks that are more stable, less likely to fail converging on a valid solution, and better able to capture more complex correlations in the input data.

The performance of LSTM and GRU networks are viewed as comparable in many practical scenarios [12]. GRU networks are simpler to implement and are computationally more efficient. For these reasons, we opted to explore the use of a GRU network for our analysis.

GRU cells have complex internal structure (Fig. 2), involving a reset gate and an update gate, with their individual activation functions, usually based on the sigmoid function, *σ*(*x*) = 1*/*(1 + *e*^−*x*^), or variations thereof. The activations of the reset gate, *r*_*t*_, and update gate, *z*_*t*_, given by

**Fig. 2.**
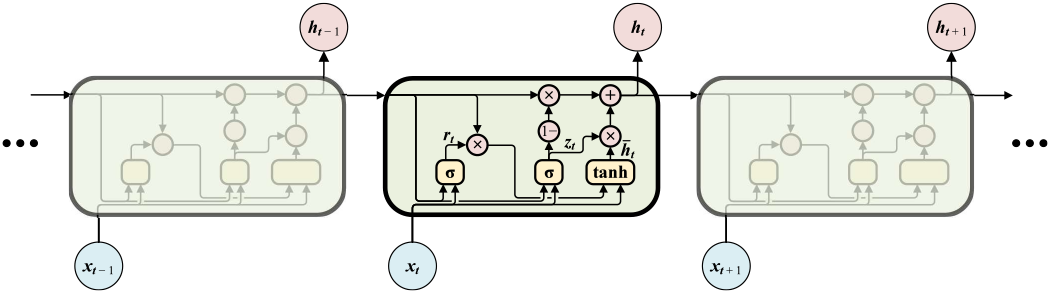
Schematic of a layer in a gated recurrent unit (GRU) network. Structurally similar to RNN cells, GRU cells have more complex internal structure involving a reset gate and an update gate, with associated additional weights.

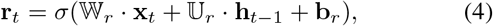

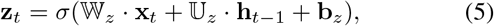

have their own respective weight matrices 𝕎_*r/z*_, 𝕌_*r/z*_ and biases **b**_*r/z*_, shared across the cells in a layer. The activation *h*_*t*_ in the GRU cell is determined as a linear interpolation of the previous activation *h*_*t*−1_ and the candidate activation 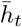,

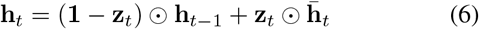

(where (⊙ is used to indicate element-wise multiplication of two vectors or two matrices of the same dimension) with 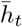 computed using

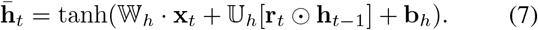

Finally,

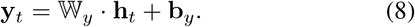

LSTM cells have similar structure, but feature a “memory”, the state of which is also propagated between cells. In most practical situations GRUs have been found to offer comparable performance with reduced complexity. Therefore, we opted to use a GRU network in this first iteration of our attempt to use machine learning to analyze MIMIC data. Similar to RNNs, GRU networks are also trained through backpropagation: after a loss function is defined, gradients are computed and steepest descent or a related method is used to update weights and transformation matrices. Given, for instance, a loss function *L* = *N*^−1^ ∑ (**ŷ**_*t*_ − **y**_*t*_)^2^, where **ŷ**_*t*_ are the expected output values and *N* is the length of the time series sequence, backpropagation is accomplished by computing the gradients

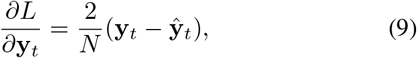

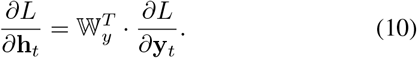

For the final timestep of a sequence, we use *δ***h**_*t*_ = *∂L/∂***h**_*t*_. For prior timesteps, as we proceed backwards, we calculate

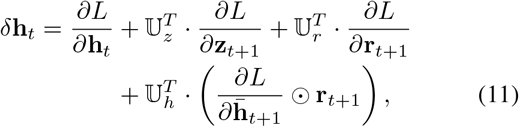

for the last layer. If the network has more than one layer, for prior layers we use instead

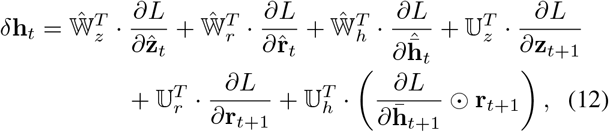

where we used a hat to indicate values taken from the subsequent layer. We then obtain the gradients

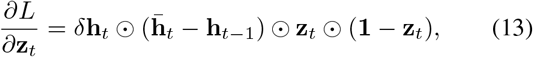

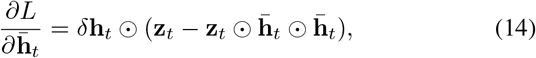

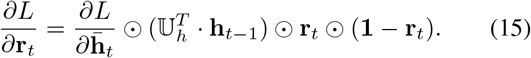

Finally, we can compute the gradients we need (*⊗* represents the outer product):

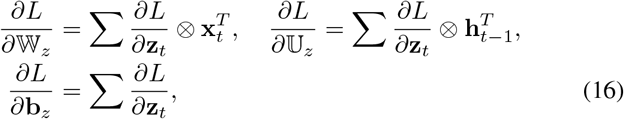

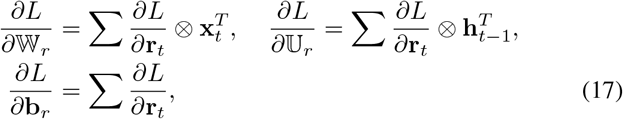

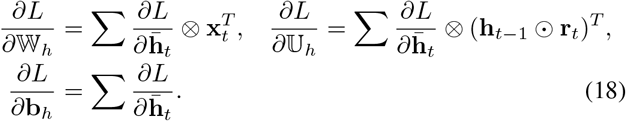

Also,

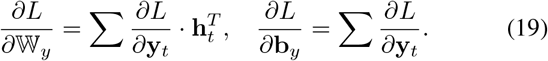

These gradients are accumulated across all cells in a layer. At last, we can update the weights and biases of the model for the next iteration:

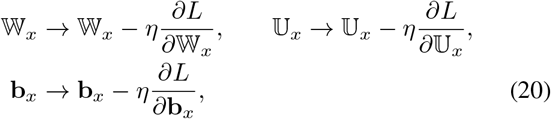

where *x* = *z, r, h, y* and *η* is the learning rate, a model hyperparameter.

However, using such a global learning rate is coarse and suboptimal, leading to slow convergence or instability. Therefore, adaptive learning rates are often employed to improve a model’s performance. One possibility is the so-called Adam optimization [13], [14], which involves establishing a set of first and second moments, **m**_*e*_ and **v**_*e*_, which are defined and updated individually for each model parameter using the equations

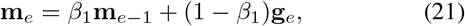

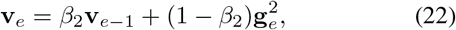

where *β*_1_ and *β*_2_ are hyperparameters that control the exponential decay rates of the moving averages for the first and second moment estimates, respectively, with *β*_1_ = 0.9 and *β*_2_ = 0.999 being typical values, and where **g**_*e*_ is the gradient of the loss function with respect to model parameter *θ*_*e*_ at training epoch *e*.

The model parameter *θ*_*e*_, then, is updated using

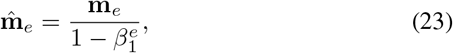

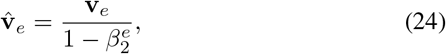

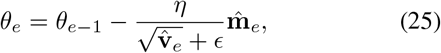

where *ϵ* is a small constant for numerical stability (a softening parameter) and the fractional expression in (25) is componentwise division.

A GRU network processes data in sequences. The length of the sequence corresponds to the number of cells in the GRU network’s input layer. Sequences may be processed independently, or the network may be constructed to maintain *statefulness*, with the initial hidden state of each sequence being the final hidden state of the preceding sequence in the temporal order of the time series data.

Both RNNs and GRU networks are capable of dealing with multiple simultaneous streams of data, in which case quantities such as **x**_*t*_ become vector-valued quantities. The conceptual foundations remain the same, except that instead of scalar operators, vector, matrix, or element-wise operators are used as appropriate to process the network.

Furthermore, it is possible to stack multiple hidden GRU layers for more complex training scenarios. The “horizontal” size of the network (length of the time sequence processed using the same set of weights and biases) and “vertical” height (number of layers) can be thought of as hyperparameters of the model.

In particular, a well-constructed GRU network with appropriate hyperparameters can learn transferable knowledge about the relationship between its inputs and outputs. For example, Figure 3 (top) demonstrates a test case in which a GRU was able to learn amplitude modulation. After being trained on an amplitude-modulated input signal for a few cycles, it was able to demodulate the subsequent signal, even when the modulation waveform and frequency differed significantly from the training data. In contrast, when the model suffers from overfitting, it memorizes instead the shape of its input, which it then erroneously attempts to apply; an example case is shown in Figure 3 (bottom).

**Fig. 3.**
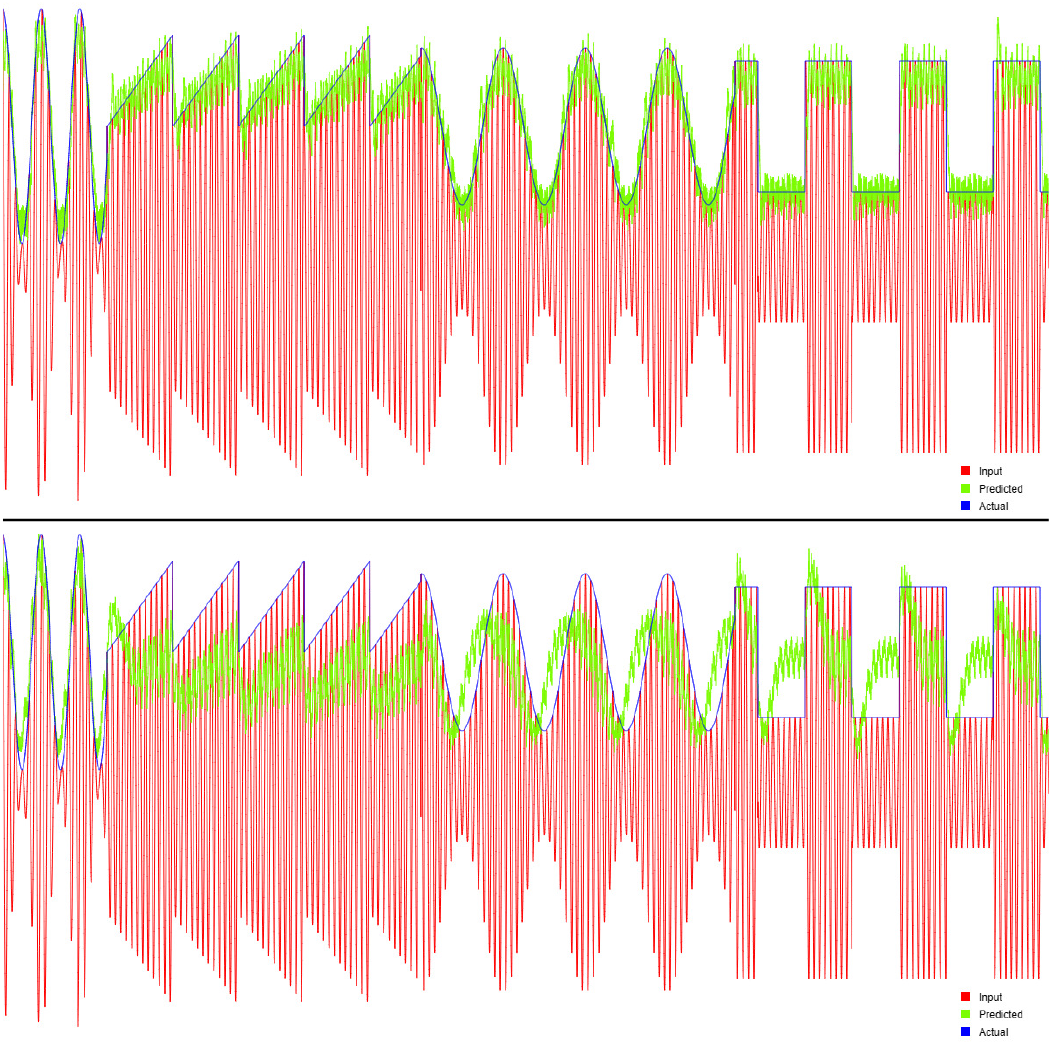
Example application of a GRU network, showing its ability to learn amplitude modulation. The network was trained on the first 10% of the data set, a carrier modulated by a sine wave modulating signal. The model was then tested using signals of different frequency and modulation. Top: The model acquired transferable knowledge that it was able to use to correctly demodulate all signals. Bottom: An overfitted model memorized the initial training data and attempted to use it inappropriately, even when the input signal was modulated differently.

## III. Adapting a GRU solution to MIMIC-IV

As part of a preliminary supplement to the MIMIC-IV data release, waveform data were released for 200 hospital visits involving 198 patients. Waveform data are presented in the FLAC (Free Lossless Audio Codec) format, a standard format for audio frequency data sets that can be easily converted into other convenient formats using off-the-shelf, open source software tools. In addition to the raw waveform data, the release also includes processed data sets, time series records with numerically interpreted values, similar (or identical) to the numerical values that might appear on the displays of intensive care instrumentation.

Our goal of developing a tool for machine learning assisted data analysis was centered around these numeric data sets. Therefore, we recap first the nature and format of the data that have been made available in this form.

Table I summarizes the columns that appear in the numerical data accompanying the MIMIC waveform data set. It is important to note, however, that not all columns are recorded for all patients. Furthermore, the data are often intermittent, or not recorded for the entire duration of a patient’s stay.

**TABLE I.**
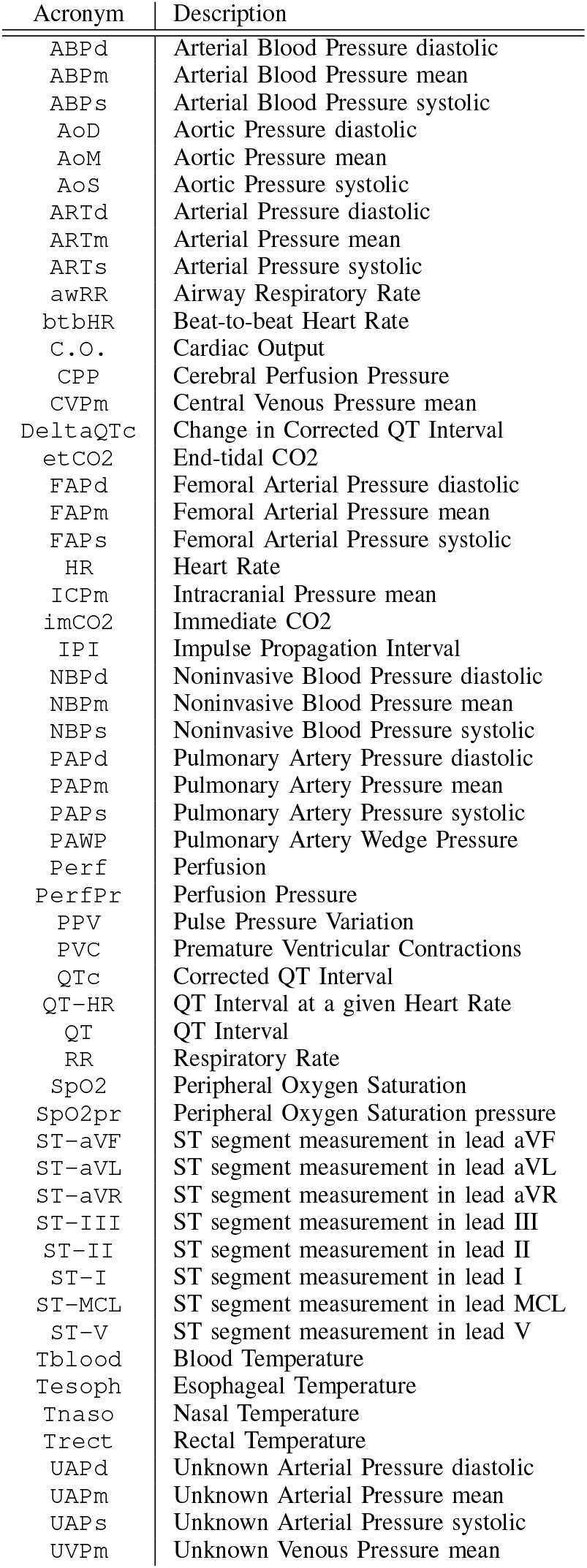
Column labels that appear in the numerical value sets associated with the preliminary (V0.1.0) MIMIC-IV waveform data release.

Nonetheless, for many patients, extended periods (several ten hours) of data are available, allowing us to use machine learning tools to discover possible nontrivial correlations. We were particularly interested in the possibility that data corresponding to non-intrusive measurements, such as heart rate or ECG, might be used to reliably estimate, perhaps even predict, values that can only be obtained by intrusive measurements, blood pressure in particular. The MIMIC-IV release, even in the form of this preliminary supplement, contains data from several dozen patients that appear to be suitable for this purpose, allowing us to proceed with constructing an appropriate machine learning tool and test the feasibility of its application.

The data are presented as time-ordered time series; however, the sampling is not necessarily done at regular intervals, and different parameters are sampled at different rates. This makes it difficult to analyze the data using machine learning algorithms. To address this problem, we opted to resample the numerical data using a Gaussian moving window algorithm.

Once the data series are resampled at uniform intervals, they can be readily used with a recurrent neural network to discover nontrivial autocorrelations within data sets, or correlations between data sets. Our specific goal was of the latter aspect: We were especially interested in finding out whether or not it would be feasible to predict the behavior of certain time series from other, parallel streams of measurement and thus anticipating the possibility that non-intrusive measurements may serve as reasonably reliable proxies for measurements of a more intrusive type. We focused our attention in particular on cases in which a neural network might be trained during a relatively benign period when the patient’s parameters are stable in order to see if the network can nonetheless predict alarming, perhaps catastrophic changes, thereby offering an opportunity to alert health care workers before a patient’s condition becomes critical.

We recognize that our analysis of a limited, early-release data set represents an approach that is strictly experimental. Our effort is aimed at exploring the feasibility and limits of extracting useful information from MIMIC waveform data sets, and determining if sufficient, discoverable correlations exist within these data sets that might one day offer clinical utility.

For this reason, transparency, ease of use, and ease of modification were our primary drivers when selecting the technology for our implementation. We opted to implement a GRU solution to analyze MIMIC-IV waveform data in a Web browser, using mostly browser-based JavaScript, with a small amount of server-side code to allow for the efficient retrieval of selected data sets.

In responding to emerging concerns about “code bloat”: the proliferation of third-party libraries of uncertain origin and dubious quality, often incorporated to accomplish relatively trivial tasks, we opted not to use third party code at all in this implementation. Initially we allowed one exception: We used the `TensorFlow.js` [15] JavaScript library for a GRU network implementation in an initial version. Eventually, however, we replaced this implementation with a “home-grown” GRU solution, which, for our purposes, actually performed better than the `TensorFlow.js` library. (The current version of our software makes the choice of library user-selectable.) The basic visual layout of our application is shown in Fig. 5. As this layout indicates, utilizing the application is done in three stages: data selection, smoothed resampling, and machine learning.

### A. Data selection

Data selection begins with the back-end code listing all available files. We collected all `.csv` (comma-separated values format) files (195 files in total in the present MIMIC pre-release) containing time series measurements in a folder. The file names serve as patient identities, which are listed in a dropdown control. The files vary greatly in size, ranging from a mere 1231 bytes to over 140 megabytes. The following brief excerpt from one of the shortest files illustrates the basic structure:

~~~
“time”,”Perf [NU]”,”Pulse (SpO2) [bpm]”,”RR [rpm]”,\
“SpO2 [%]”,”btbHR [bpm]”
7552,,,28,,
8576,,,27,,
9600,,,26,,
10624,,,27,,
11648,,,28,,
12672,3.2,25,28,95.5,
13696,3.2,25,28,95.5,
13710,,,,,181.27
14720,3.4,25,30,93.2,
13710,,,,,181.27
[…]
~~~

The actual columns vary from file to file. It is important to note that there are missing data. For example, most rows in most files have values only for select columns. In addition, some measurements are simply performed less frequently. Other measurements may be altogether absent for significant durations of a patient’s visit.

Each data file is accompanied by a header file in the WFDB format^1^. The header file contains some relevant information about the data content. For the example above, the corresponding header file contains the following lines:

~~~
#wfdb 10.7
86239129/2 3 62.4725/999.56 4560 10:40:05.181 18/6/2123
86239129_0000 0
86239129_0001 4560
# subject_id 19477300
# hadm_id 29371273
~~~

Of particular importance to us is the *counter frequency* field, in this case the value of 999.56 in the second line of this file. The reciprocal of this number basically amounts to specifying the units of time, i.e., the first column in the `.csv` file of measurements. Translating the time column accordingly, it tells us that the first measurement in the preceding `.csv` listing took place at timestamp 7552, which is to say, at 7552*/*999.56 *≃* 7.56 seconds.

The header file also records the subject (patient) identifier. The MIMIC data release contains basic (anonymized) patient information elsewhere. Unfortunately, this information is not available for all patients in the present waveform data prerelease. This, along with the very small set of files presently available, precludes us from attempting to train models that may learn relationships between measured parameters that would be transferable between, e.g., patients of similar age and gender.

After the user picks a patient, the set of available columns is shown in the user interface, allowing the user to pick a desired subset and then use the Get Data button to load that set. The data is then loaded into a variable in the user’s Web browser, where it will be processed.

### B. Resampling

Data resampling takes place, as discussed earlier, using a Gaussian window function with user-configurable parameters. Given a time series data set sampled at irregular intervals, yielding values in the form *v*_*i*_(*t*_*i*_), we resample the data using

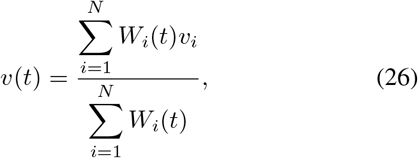

where we define the Gaussian weight function as

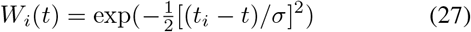

and where we treat the standard deviation *σ* as a hyperparameter. In the practical implementation, rather than resampling the entire data set (which can be time consuming if there are a large number of data points, most of which would not contribute to the sum) we stop the sampling when the Gaussian weight function vanishes. This allows us to rapidly resample input data.

Choosing a large *σ* will smooth the data set; conversely, choosing a *σ* that it too small may introduce unnecessary noise, depending on the sampling rate of the original data set and the desired sampling interval, which is also treated as a hyperparameter.

Concerning optimal values for the desired sampling interval *δt* and *σ*, we obtained good results by choosing *δt* = *σ* =𝒪 (10 s). Actual choices should reflect the nature of the available data, the number of data points present in the data set, and other model parameters that will be discussed below. In other words, *δt* and *σ* must be treated as model hyperparameters, with the choice of their values guided by the system’s ability to model the data set accurately.

### C. Data analysis using a GRU network

With the time series data sets resampled, we are in the position of applying our GRU network to analyze the data set. Our goal is to check if it is indeed possible, at least in some cases, to use the time series values of certain measurements as predictors of other time series. We are in particular interested in being able to predict values of blood pressure from non-intrusive measurements, such as the patient’s heart or respiratory rate.

The GRU network that we implement must maintain statefulness: subsequent sequences of data represent a progressive time series, and statefulness is required to correctly capture longer-term trends in the data.

The GRU network has many hyperparameters. Choosing the right set of hyperparameters is essential if we hope to obtain consistent, reliable predictions. The wrong hyperparameters can yield invalid or overfitted solutions.

Neural networks make extensive use of random number generation (RNG), as a means of initializing the network’s weights and biases. In order to generate reproducible results, we opted to control the implementation of the RNG to ensure that results can be consistently replicated. The seed of our RNG is one of the model hyperparameters that can be specified by the user. We note that if machine learning of this nature is ever going to be deployed in a clinical environment, reproducibility might be essential both to guarantee the integrity of research results and also for ethical and legal reasons. Therefore, we stress that explicit control over RNG behavior is essential.

With these considerations in mind, we opted to expose several hyperparameters through the user interface. In addition to specifying the dependent column and the RNG seed, the following parameters can be controlled by the user:

- Parameters characterizing the model size, including the length of a sequence (window size) that is processed by the GRU network in one pass, the number of hidden layers in the GRU network, and the hidden unit size that characterizes the model dimensions for each layer;
- Parameters that control the training such as the maximum number of learning epochs, percentage at the beginning of the data set that is used for training, and the learning rate;
- Parameters that control the adaptive training rate, managed by an Adam optimizer (if selected);
- Parameters that control early stopping, including the model patience (number of epochs with no improvement) and the percentage of the training data set that is used for model validation.

The run of the GRU network can be monitored as the value of the loss function is displayed after the completion of each training epoch. If the model drops the learning rate or stops, that is also indicated. When the run is complete, the user interface displays a plot comparing the predicted values of the dependent column against actual values. A *χ*^2^ per degree of freedom result is also displayed, as a metric characterizing the goodness of the fit.

## IV. Preliminary results

With the model complete, we ran numerous tests on a several of the available patient data files. It is unfortunate that many of the patient data files in this preliminary release contain only a few columns of data, that the data sets are often incomplete, and numerous outliers are present (see Fig. 4). As a result, out of the nearly 200 data files only a small number of the files are of sufficient quality to test our neural network’s ability to predict values.

**Fig. 4.**
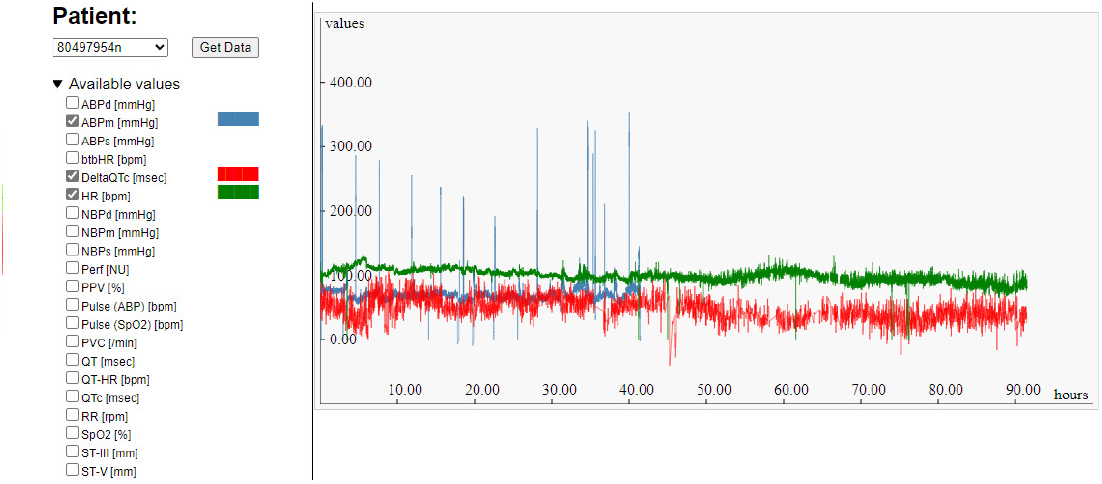
A typical example from the MIMIC waveform database. Note how the mean arterial blood pressure reading is contaminated with numerous outliers and ends prematurely, less than halfway into this patient’s stay.

Despite these limitations, we observe some encouraging results. One of the best examples is shown in Fig. 5. The first panel of the user interface shows the raw data, with three columns selected: mean arterial blood pressure (`ABPm`), heart rate (`HR`) and respiratory rate (`RR`). About 22 hours into his stay at the emergency room, the patient clearly suffered a life-threatening incident, with heart rate and respiratory rate first increasing slightly and then dropping, even as the patient’s blood pressure collapsed. In this case, the blood pressure was continuously monitored, but could the collapse in the blood pressure have been predicted based solely on the variations of the monitored heart and respiratory rates?

**Fig. 5.**
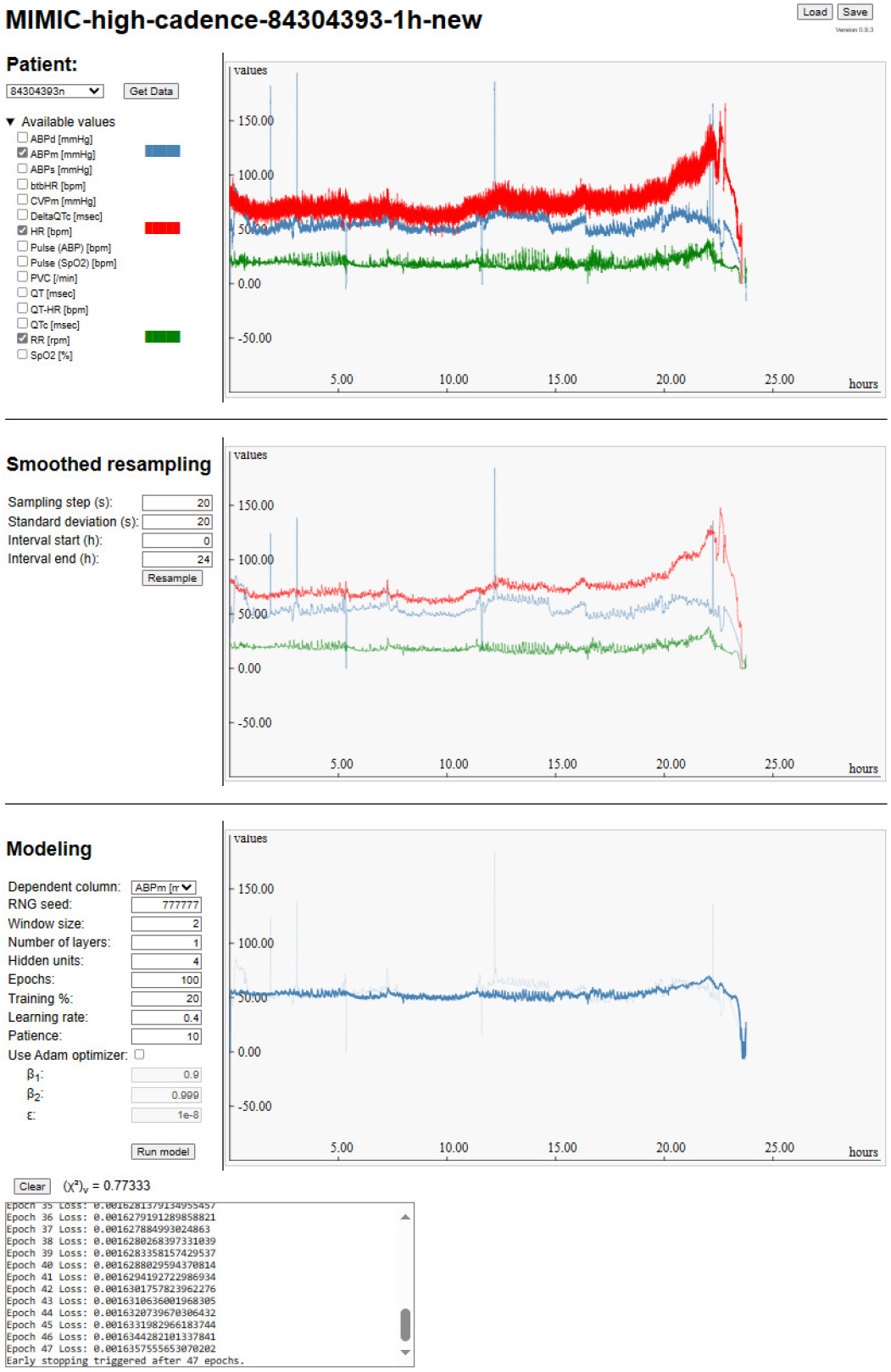
The MIMIC data analysis software user interface, showing three panels: Input data, smoothed and resampled data, and fit results.

Indeed, after the data were smoothed (second panel in Fig. 5), our learning model, trained on the first 20% of the patient’s data (roughly the first five hours of this patient’s IR stay), was able to predict correctly, from heart rate and respiratory rate, the observed precipitous drop in blood pressure. Moreover, throughout the patient’s stay, the model accurately predicted the mean blood pressure, with an impressive (*χ*^2^)_*ν*_ = 0.77333.

While the above results are promising, there are limitations. Presently, the model remains quite sensitive to even small changes in its hyperparameters, including the RNG seed. This indicates that the model is not stable. Instabilities emerge in a variety of ways. The model may, instead of accurately predicting the dependent column, “lock in” to a flat line average. Conversely, the model’s predictions may be excessively noisy. In some cases, the model correctly followed mild changes in the dependent column but large swings, such as the one near the end of this particular patient’s stay shown in Fig. 5, were modeled incorrectly. Clearly, these issues are significant, and before our results can have any practical application, we must explore and understand this behavior in depth and identify root causes.

For the time being, we found the following general features:

- The model tends to work better with a fixed learning rate. The Adam optimizer may slightly improve stability, but the resulting fits were generally inferior;
- A very short window size yields best results. Longer window sizes resulted in overfitting, often manifesting as the aforementioned “flat line” behavior;
- A larger number of hidden units also resulted in overfitting;
- Similarly, the model did not perform well with more than one hidden layer.

As an exercise in code validation, we also ran similar cases using the `TensorFlow.js` code base, and obtained similar results and insight. On the other hand, we were very pleased to see that the model had good performance, even with a custom GRU implementation that lacks any numerical performance optimization.

## V. Summary and plans

Using the extensive MIMIC-IV database of emergency room data, we attempted to build a data analysis tool, employing machine learning technology. Our goal was to find out if it is possible to construct a real-time data analysis capability that could be used to predict intrusive measurements from other, less-intrusive time series data sets. As a scenario, we considered an emergency room patient who faces a life-threatening condition due to circulatory collapse. This would be detected if the patient’s blood pressure was monitored, but such monitoring is intrusive and can even be painful. On the other hand, less intrusive measurements, such as the patient’s heart rate, respiratory rate, or even real-time ECG measurements may be readily available. A machine learning based system that provides early warning to health care professionals may, in such situations, offer a life-saving capability.

To this end, we looked specifically at a novel subset of the MIMIC-IV database, the preliminary release of waveform data, and actual recordings of measurements taken by emergency room instrumentation. We have not attempted to convert waveform data directly into numerical data sets; this conversion was already performed as part of the MIMIC-IV waveform preliminary data release, so we had ready access to high-cadence measurements.

The major challenge that limited the scope of our current investigation was the uneven data quality in this data release. As the data was collected across a range of patients and facilities, often very different values were measured. The measurements are sometimes interrupted or prematurely terminated. Furthermore, the data itself are noisy, often with significant outliers or obvious systematic errors.

Nonetheless, we were able to identify a few cases of “clean” data sets, including data sets that characterized patients who had an unfortunate outcome (either death or a life-threatening emergency). Such data sets were of special interest to us, since a crucial test would be the ability to train a neural net to indicate when such a life-threatening condition develops even in the absence of intrusive measurements.

Use of the MIMIC-IV waveform data prerelease, despite its limited number of patient records, was sufficient for our purposes as we did not aim to construct a pretrained model. A pretrained model may be possible with a much larger data set, especially if the records can be grouped by demographic data and patient history.

For the time being, however, our investigation had a more limited objective: Namely, to establish the feasibility that a model, which may be trained in real time during the initial few hours of a patient’s visit (or possibly, during a prior visit, if patient records are kept), can reliably predict an emerging life-threatening condition (specifically, collapsing blood pressure) from non-intrusive measurement data. Such a solution may eventually become a common emergency room tool, helping to ensure that patients in critical condition receive emergency assistance in a timely manner.

Our findings so far are encouraging but preliminary. We have indeed found cases where such predictions were possible. On the other hand, we found that the machine learning model that we employed, a GRU (Gated Recurrent Unit) network, is not always stable and its success had undue dependence on the model’s hyperparameters.

The software that we developed for this analysis can be run in a standard Web browser. It has been made available under a public, open source license^2^. We shall continue to improve our solution and seek to obtain extended data sets, either in the form of an upcoming, more complete MIMIC-IV waveform database or by using similar data from the MIMIC-III clinical database released earlier. We hope that this will allow us to validate our expectations and aid us in developing a solution that, at some future point in time, may become sufficiently robust and reliable for clinical use. Our results will be reported elsewhere as they become available.

## Data Availability

This study used the publicly available MIMIC-IV Clinical Database and MIMIC-IV Waveform Database, which are hosted on PhysioNet (https://physionet.org/). Access to the data requires credentialed approval through PhysioNet's data use agreement. No new patient data were generated in this study. The analysis code developed for this work is available on GitHub at https://github.com/vttoth/MIMIC-WF.

## Acknowledgments

The research reported in this paper was supported by CRISP (Center for Research on Integrated Systems Platform) of the Department of Systems and Computer Engineering at Carleton University. VTT acknowledges the generous support of Plamen Vasilev and other Patreon patrons.

## Footnotes

1 https://physionet.org/physiotools/wag/header-5.htm

2 https://github.com/vttoth/MIMIC-WF

## Notes

### Competing Interest Statement

The authors have declared no competing interest.

### Author Declarations

This study used the de-identified MIMIC-IV waveform database (PhysioNet). Because the dataset is publicly available in de-identified form, the study was exempt from institutional review board approval.

